# Modeling COVID-19 epidemic trends and health system needs leading to projections for developing countries: a case study of Thailand

**DOI:** 10.1101/2021.11.05.21265992

**Authors:** Nawaphan Metchanun, Christian Borgemeister, Caitlin Bever, David Galick

## Abstract

Thailand was the first country outside China to report a COVID-19 case but had a mild impact from the outbreak especially at the beginning of the pandemic. This study systematically investigates the evolution of the COVID-19 epidemic in Thailand from January 2020 to March 2021 to uncover the COVID-19 situation in the country. By modeling all health districts throughout the country, the study found that COVID-19 contributed to an increase in excess deaths and that COVID-19 deaths might be underreported. There was a lag time in ramping up testing although testing is key to control the disease. The estimated total number of beds required by COVID-19 seems low, but it may not ensure the capacity to take care of critical cases that required ICU beds, specific medical equipment, and trained medical staff.

## Introduction

SAR-CoV-2 has infected more than 200 million people and taken away more than 4.5 million lives worldwide (1). The Coronavirus pandemic has exhausted and overwhelmed health systems, especially in low- and middle-income countries (LMICs) where the health resources are fundamentally limited (2). Even with the presence of current vaccines that have been urgently developed, countries continue to suffer from high infection and mutations that complicate the battle against the disease. Epidemiological research has become pivotal in guiding public health strategies. In particular mathematical modeling has proved to be the highly sought-after approach to synthesize existing information about a disease and to systematically investigate hypotheses to identify control measures that are likely to have the most significant impact (3). In this work, we investigate the trends and the impact of the COVID-19 epidemic on health resources in LMICs using Thailand as a study area. We use mathematical models to investigate potential key contributors to the disease patterns.

Thailand reported its first case of coronavirus on January 13, 2020, becoming the first country outside China to do so (2). Despite its proximity to China, Thailand reported low COVID-19 fatalities, with under 1,000 new infections per day and less than 100 cumulative deaths out of a population of nearly 70 million from the beginning of the outbreak to March 2021(3,4). Even though Thailand seemed to experience a mild impact of COVID for over a year, the country has since faced a surge of new cases and become one of 30 countries with a high number of confirmed COVID-19 cases to date (5). COVID-19 has strained the health system in the country under slow vaccine roll-out with less than 10% of the population fully vaccinated as of August 25, 2021 (4). This study systematically investigates the evolution of the COVID-19 epidemic in Thailand from January 2020 to March 2021, before the latest surge began, to systemically understand how the country could escape the worst of the pandemic for over a year. We model all thirteen health districts of Thailand separately and analyze the epidemic trends nationally.

## Materials and methods

We use an extended compartmental model framework (6) to investigate COVID-19 epidemic trends and health systems needs in Thailand. The model framework consists of four stages: susceptible, exposed, infected, and recovered, and focuses on forecasting epidemic trends and hospital needs. It includes 22 state variables, incorporating severity in the infected stage: asymptomatic, pre-symptomatic, both mild and severe symptomatic, hospitalized, and critical. We use tests per day to model the detection of symptomatic and asymptomatic infections. Death is also included as an outcome of infection. The study covers all of Thailand, using the thirteen health districts as simulation areas (Figure 1). We simulate each health district, each of which generally has a total population between 3-6 million people (7) (Supplementary 1). Health district number 13 covers Bangkok, which has a population of around 8.3 million, making it the most populated health district (7).

**Figure 1.**
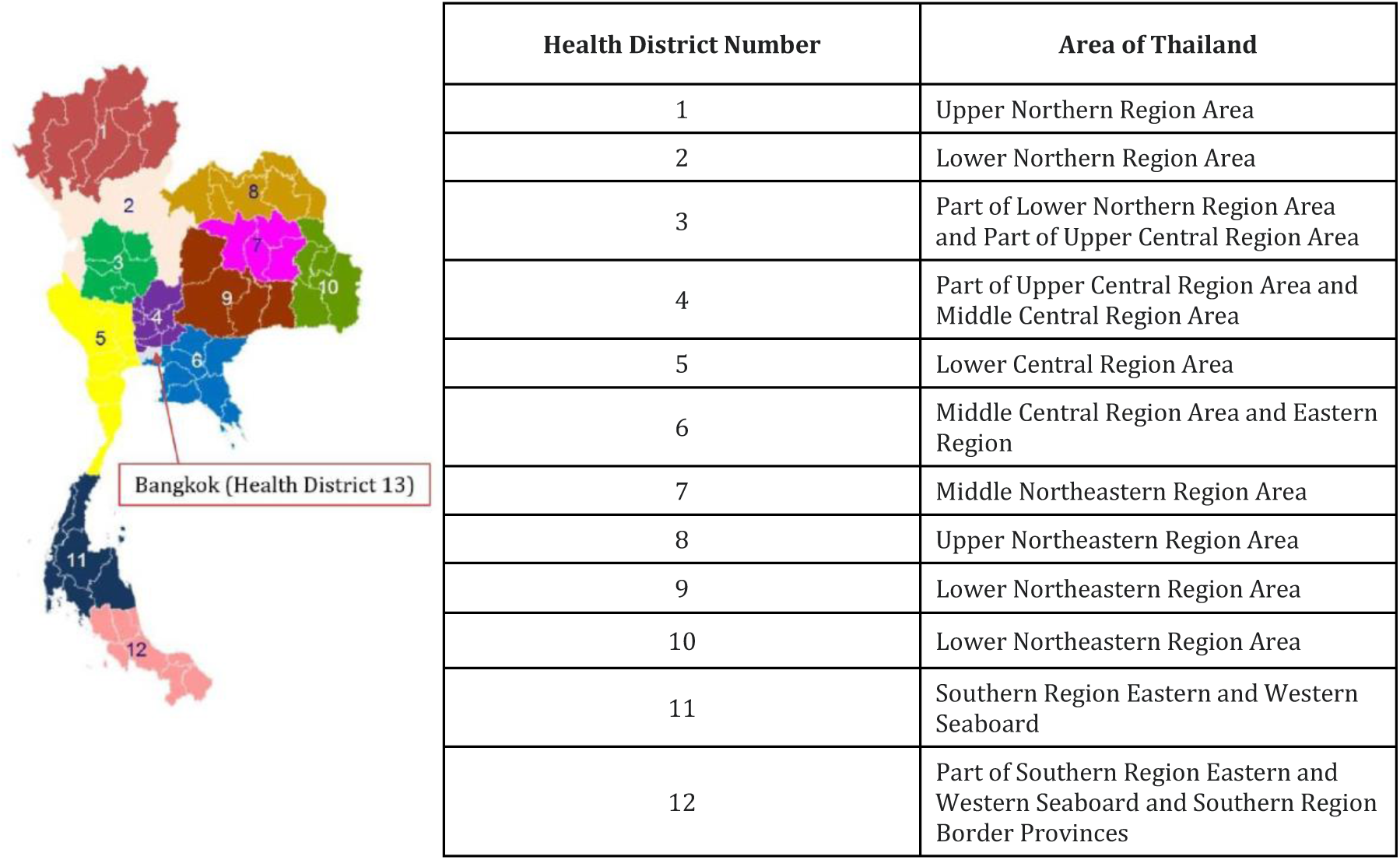
Study area includes all 13 health districts of Thailand

The model is fitted to the number of cases and deaths per province reported from January 12, 2020, to March 4, 2021, as reported by Thailand’s Ministry of Public Health (MoPH) (8). We identify the ranges of time-varying transmission rates for each health district. In calibration, all simulations start on February 1, 2020, and run for 365 days. The study parameter ranges of each health district are taken from the best-fitted calibration after 5 iterations, each with 64 simulations for a total of 320 simulations per calibration. Parameters applied in the model are derived from the literature and open data sources (6,9,10) (see the list of parameter values applied in the model in Supplementary 2). We extract the weekly COVID-19 tests performed in each health district reported by the Department of Medical Sciences, MoPH, from April 4, 2020, to February 28, 2021, and assume the test sensitivity at 0.95 based on information from the literature (11–13). We calculate the excess deaths using the monthly number of deaths per province from January 2015 to March 2021 and estimate the bed capacity of each health district using the data of hospital and medical establishments with beds by province from the year 2015 to 2019 reported by Thailand’s National Statistical Office (14). The dates of changes in the transmission rates are based on the community mobility data (15) combined with the national restriction easing plans (16,17) (Supplementary 3). Once we have the fitted time-varying transmission rate, we run 100 simulations of the 370-day period per scenario for each health district. We then analyze simulation outputs including transmission rate, the number of COVID-19 cases and deaths, hospital and intensive care unit (ICU) beds required for COVID-19 patients.

## Results

We observed a substantial discrepancy between reported COVID-19 deaths and excess deaths in most health regions (Figure 3), indicating that COVID-19 has contributed to a substantial increase in mortality. Overall, it is also possible that the burden of COVID-19 has been vastly underreported in areas outside Bangkok, except Health District 6, 11, and 12. The number of excess deaths is lower than reported deaths from COVID-19 in Bangkok, as the lockdowns might contribute to fewer deaths in the area. Note that excess deaths were not used in model fitting since excess deaths were available only at a monthly temporal resolution, and may include other causes of deaths while the model is estimating deaths from COVID-19.

**Figure 2.**
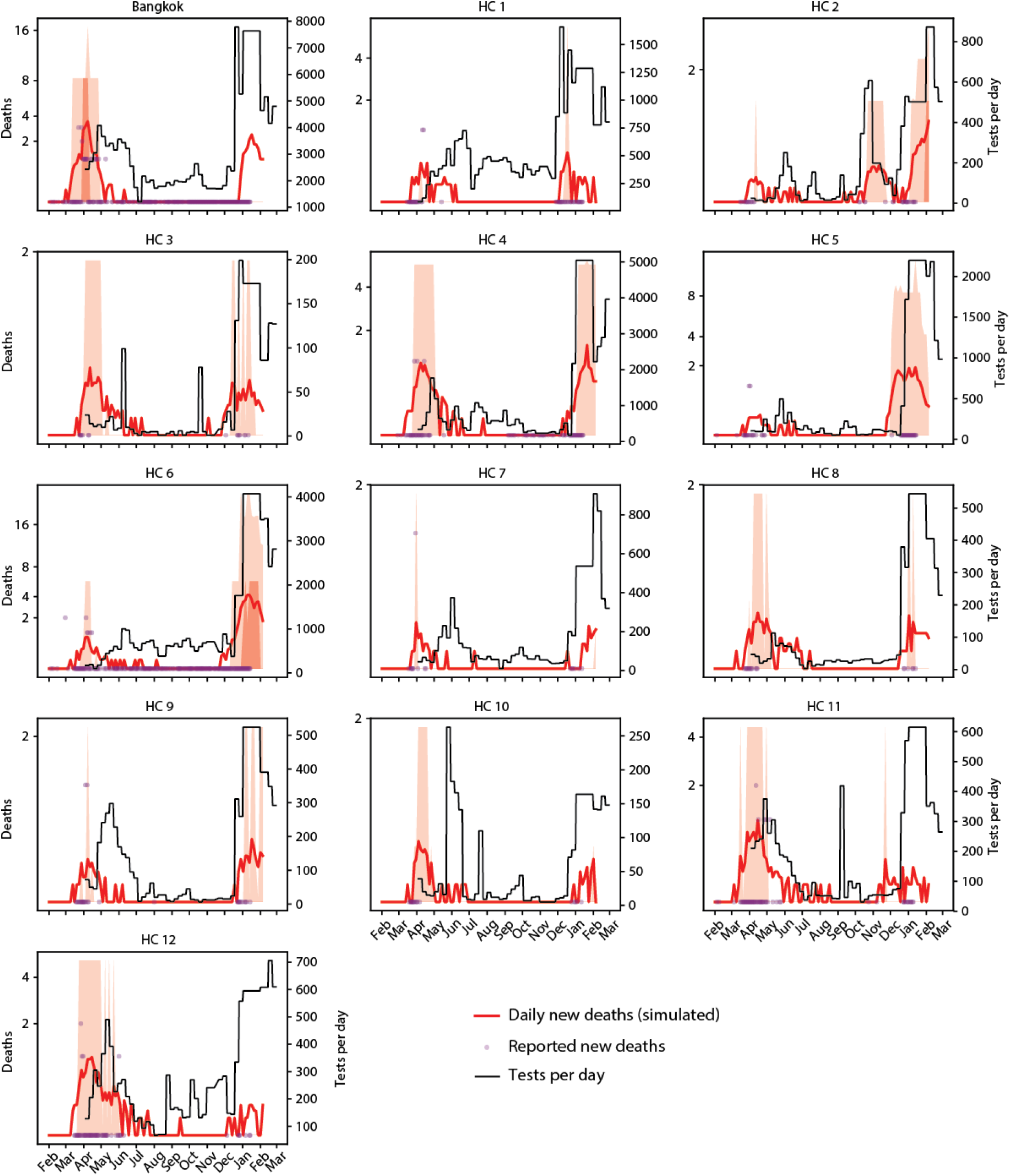
Outputs of new detected COVID-19 deaths from simulations in red, new reported COVID-19 deaths per day from Thailand’s MoPH data in purple dots and reported testing rate in black lines. Shaded areas indicate 50% and 90% predicted intervals, more intense and less intense shades accordingly, from 100 realizations.

**Figure 3.**
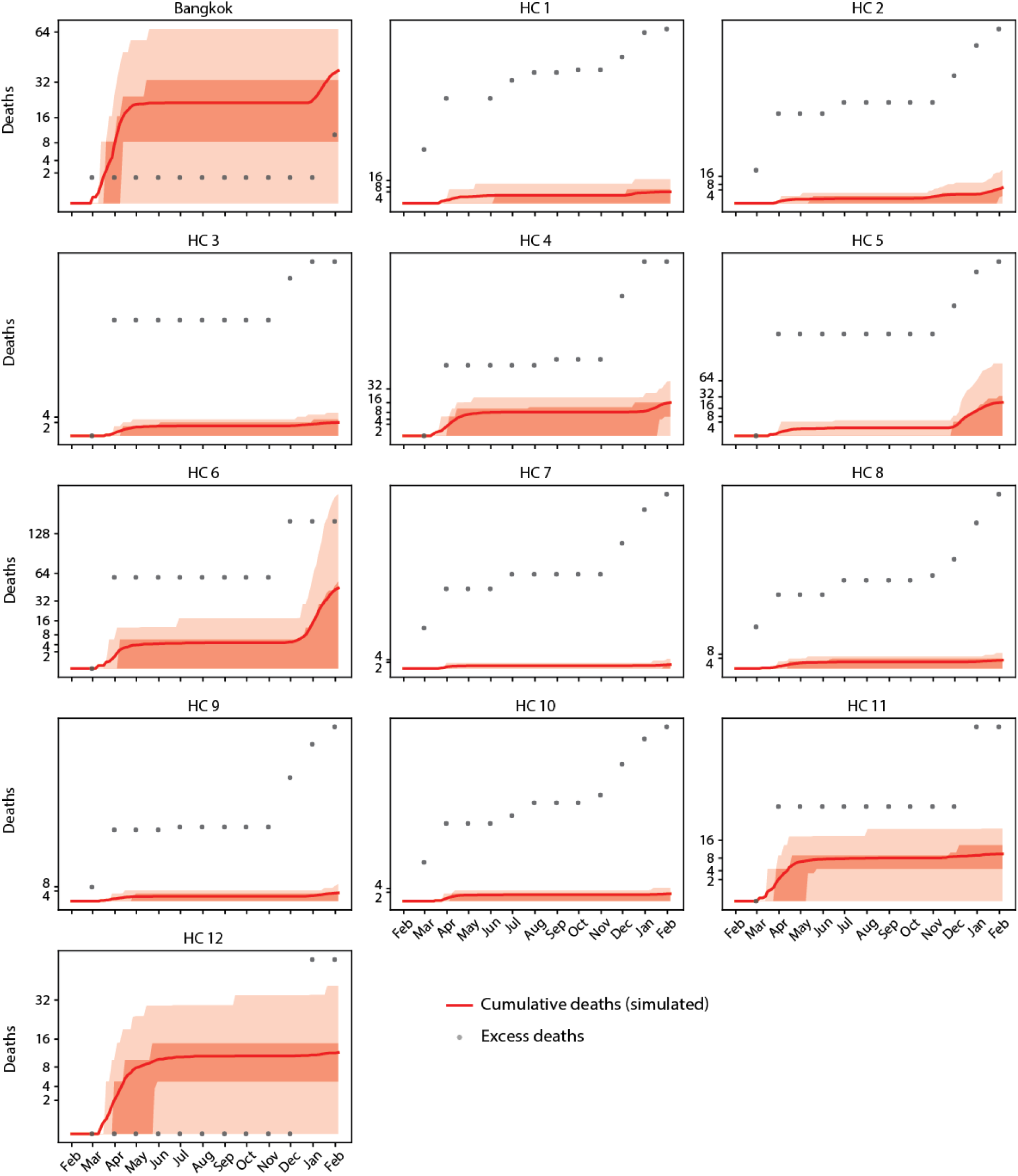
Outputs of cumulative deaths from simulations in red and cumulative excess deaths per month estimated from excess deaths estimated from all-cause deaths reported by the National Statistical Office of Thailand in black dots. Shaded areas indicate 50% and 90% predicted intervals, more intense and less intense shades accordingly, from 100 realizations.

Models fitted to daily reported COVID-10 cases and deaths provided reasonably good fits to the data and were able to explain the multiple surges observed in Thailand (Figures 2 and 5). Fitting the model to reported deaths was particularly difficult given the small number of deaths and a smaller simulated population than the true population. As a result, in some cases the model over-predicted deaths in some cases. The dates of changes in transmission rate indicate that the first wave started around the second week of March in all areas (Table S4 in Supplementary), followed by a strong reduction in transmission sustained throughout most of the year until the second wave started in October 2020 for most health districts except Health District 2 where the similar pattern was observed but the transmission rate went down after a very short while.

**Figure 4.**
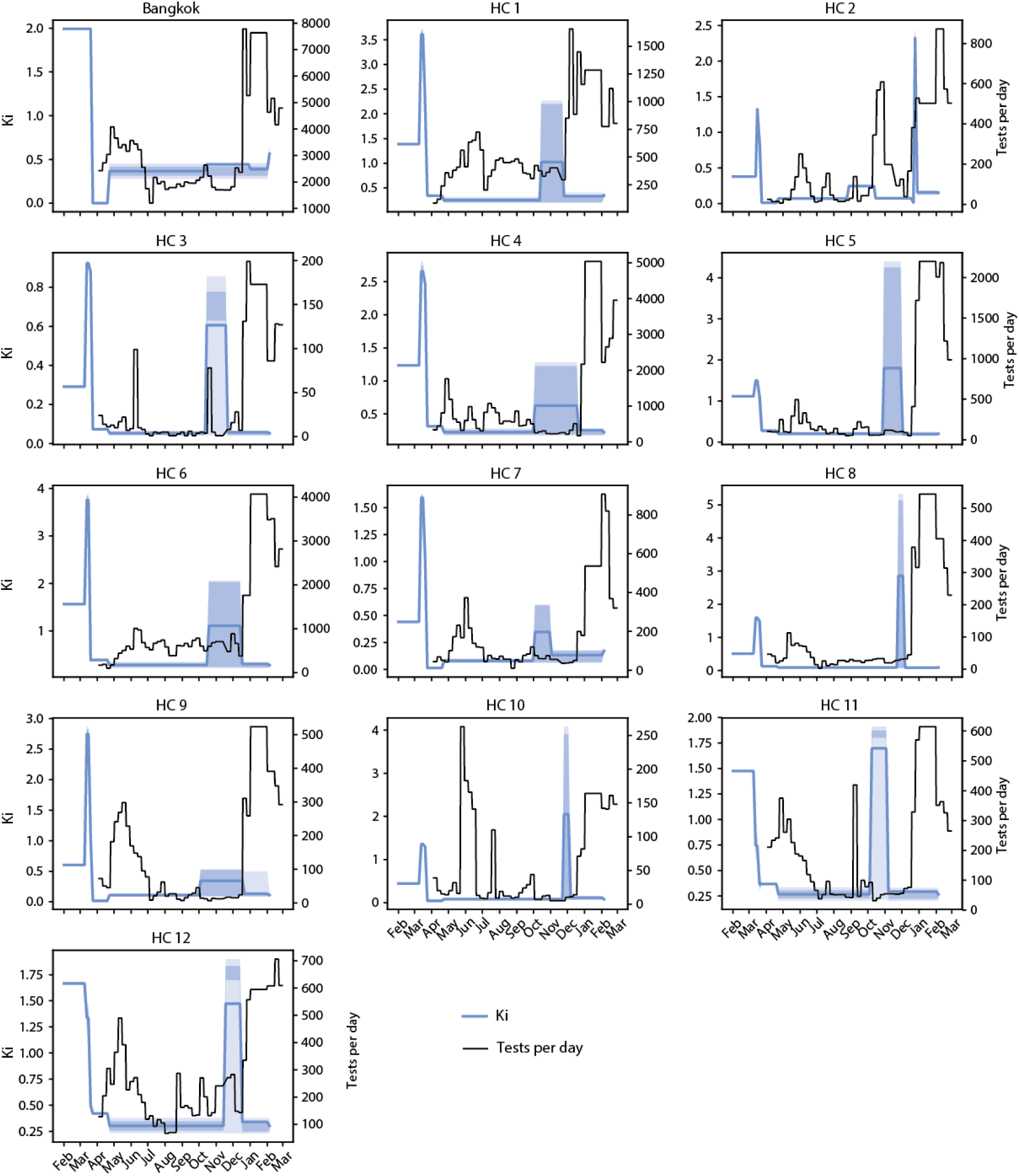
COVID-19 transmission rates estimated from simulations in blue and reported tests per day from the data in black lines. Shaded areas indicate 50% and 90% predicted intervals, more intense and less intense shades accordingly, from 100 realizations.

**Figure 5.**
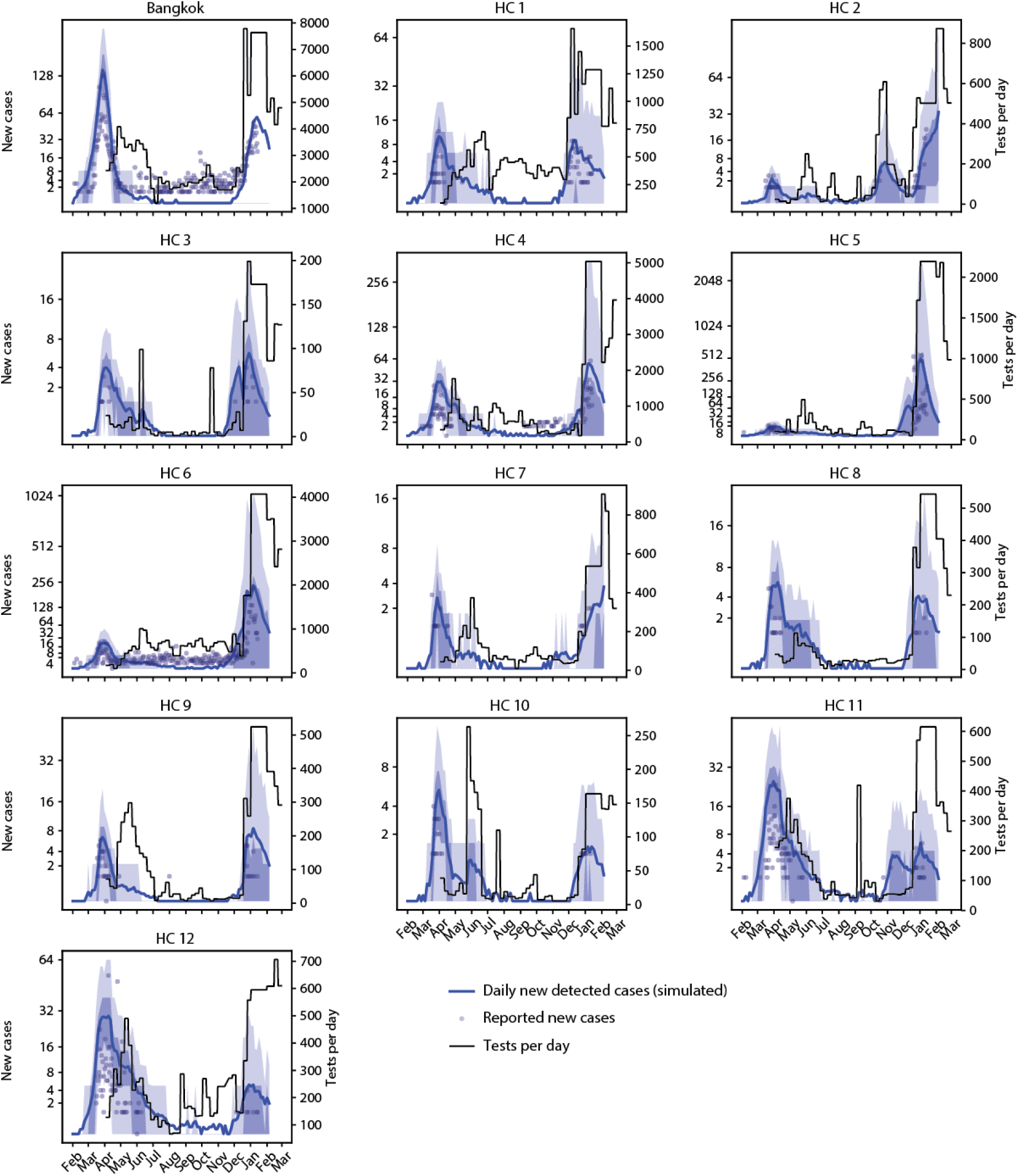
Outputs of daily detected COVID-19 cases from simulations in blue, reported COVID-19 cases per day from the data in grey dots and reported testing rate in black lines. Shaded areas indicate 50% and 90% predicted intervals, more intense and less intense shades accordingly, from 100 realizations.

The simulation results also indicate that the testing has been used overwhelmingly as a reactive intervention. Testing rates increased only after transmission rates increased in all areas (Figure 4), resulting in a lag time from one to three months from the start of the increased transmission before the testing ramp up. In the first wave, the lag between increased transmission and testing ramp-up was two to four months. For the second wave, the response time was quicker, with a lag time of approximately 15 days on average compared to around 30-60 days during the first wave.

The number of beds and ICU beds needed for COVID-19 patients estimated from the simulations is shown in Figure 6. Many of the hospitalized patients are likely to develop more critical symptoms thus further requiring critical care in the ICU. The estimated beds required are on average < 1 percent of the available beds in each health district (Table 1). It is, however, worth noting that half of the hospitalized COVID-19 cases would become critical and require ICU beds, ventilators, and trained medical staff that are not all available for all beds reported by MoPH.

**Table 1.** Percentages of estimated bed required by health district

| **Health Region** | **%Estimated beds required**<br>**(maximum/day)** |
| --- | --- |
| Bangkok (HC13) | 0.14 |
| HC1 | 0.01 |
| HC2 | 0.10 |
| HC3 | 0.02 |
| HC4 | 0.08 |
| HC5 | 0.17 |
| HC6 | 0.34 |
| HC7 | 0.01 |
| HC8 | 0.02 |
| HC9 | 0.02 |
| HC10 | 0.02 |
| HC11 | 0.07 |
| HC12 | 0.09 |

**Figure 6.**
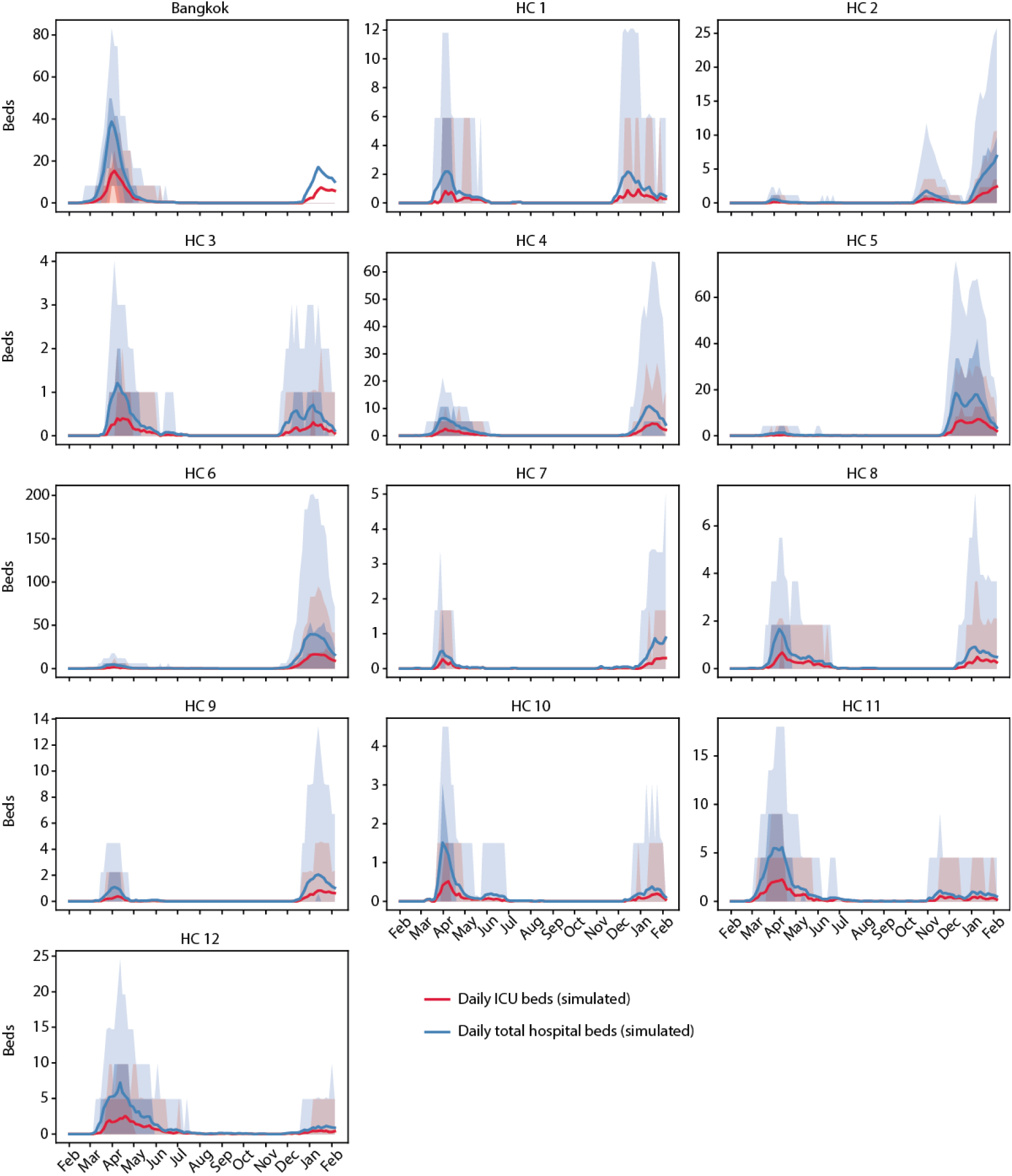
Compared the total number of beds with ICU beds required for COVID-19 patients from simulations Estimated daily total number of beds (in blue) and ICU beds (in red) required for COVID-19 patients from simulations. Shaded areas indicate 50% and 90% predicted intervals, more intense and less intense shades accordingly, from 100 realizations.

## Discussion

From our estimation, COVID-19 contributed to an increase in excess deaths and we found that COVID-19 deaths might be underreported and there was a lag time in ramping up testing. Even though the estimated total number of beds required by COVID-19 seems low, it may not cover all COVID-19 cases, especially the critical ones, as they required ICU beds, specific medical equipment, and trained medical staff.

The large increases in estimated excess deaths line up temporally with the first and second waves of confirmed COVID-19 cases and deaths. It is then reasonable to assume that these increased excess deaths were largely caused either directly or indirectly by COVID-19. Thus the real burden from the outbreak may have been substantially higher than reported. Meanwhile, the overall COVID-19 measures probably reduced all-cause mortality, excluding those caused by COVID-19. The results from Bangkok corroborate this, as there were more reported confirmed COVID-19 deaths than excess deaths, suggesting an overall reduction in all-cause mortality. This means excess deaths data may underestimate the effects of COVID-19. In Thailand. excess deaths are at least indirectly attributable to COVID-19 (Figure 3) but were not reported as COVID-19 deaths. As we see in Bangkok, all-cause mortality (minus COVID-19) was lower than excess deaths due to lockdowns and containment measures.

An important finding of the simulations was the importance of testing, aligning with the WHO’s and OECD’s recommendations that testing is key to COVID-19 outbreak management (18,19). In Thailand, once testing increased transmission levels started to drop, but there was a lag time especially at the beginning of the outbreak before the testing capacity started to catch up, causing sizeable surges. Sustained higher levels of testing might have had limited the outbreak, but the testing rate in Thailand has been fluctuating and overwhelmingly reactive. The reductions in transmission can be seen as a combined outcome of the increased detection, containment measures, and increase in awareness in the population. Additionally, the fitted changes in transmission cast doubt on whether the 5-phase easing plan was appropriate since transmission rebounded around one month after the completion of the easing plan.

The estimated beds required were low, but half of the hospitalized COVID-19 cases would become critical and require ICU beds and other medical equipment that required trained medical staff. Our study shows that not being complacent in testing even when the transmission is low might result in better control of the outbreak even when the transmission is low. This highlights the importance of testing aligning with WHO’s recommendations (19). It is then not a recommended practice of easing restrictions without maintaining testing amidst a low vaccination rate. It is worth noting that the findings are speculative to some extent since this study did not model possible alternatives of reopening with a high testing rate. Data indicated that Thailand had focused on COVID-19 testing of incoming travelers and was not as proactive in detecting local outbreaks (20). The main testing method available in Thailand is Real-time Polymerase chain reaction (RT PCR) that took 24-48 hours to deliver the test results (20). This waiting time can contribute to the delay in case management especially when people who get tested are not quarantined. The authority advises home quarantine during this waiting period but there is no monitoring system in place to efficiently monitor these cases. Our simulation results support WHO’s recommendation that the sooner the testing can be initiated especially among vulnerable populations and risk groups, more testing and early detection help reduce transmission and the number of cases that lead to fewer severe cases and deaths (19).

Our model helps fill the gap of understanding the size of the COVID-19 outbreak and can be used to monitor epidemic trends in the areas where data are limited. The ability to estimate and monitor the epidemic trends and health system needs are valuable for immediate measures to prevent the further spread of the disease and contribute greatly to the health system resource management in the areas.

## Data Availability

All data produced are available online at

https://github.com/NaniMet/covid-Th

https://github.com/numalariamodeling/covid-chicago

https://github.com/InstituteforDiseaseModeling/dtk-tools

## General

The authors thank the Institute for Disease Modeling and, in particular, Edward Wenger for their generous technical support and resource sharing.

## Author contributions

Conceptualization and experimental design: NM and DG. Experimental work: NM and DG. Data interpretation and analysis: NM and DG. Supervision: CB. Writing original draft: NM and DG. Writing – review and editing: NM, DG, CB, and CB.

## Data and materials availability

Codes on GitHub

COVID-Thailand: https://github.com/NaniMet/covid-Th

COVID-Chicago: https://github.com/numalariamodeling/covid-chicago

dtk-tools: https://github.com/InstituteforDiseaseModeling/dtk-tools

