## Supplementary file for "Modeling COVID-19 epidemic trends and health system needs leading to projections for developing countries: a case study of Thailand"

Table S1 Details of health districts of Thailand (8).

| Health District Number | Area of Thailand | Area Code | Provinces |
|---|---|---|---|
| 1 | Upper Northern Region Area 1 | 15 | Chiang Mai, Mae Hong Son, Lampang, Lamphun |
| | Upper Northern Region Area 2 | 16 | Chiang Rai, Nan, Phayao, Phrae |
| 2 | Lower Northern Region Area 1 | 17 | Tak, Phitsanulok, Phetchabun, Sukhothai, Uttaradit |
| 3 | Lower Northern Region Area 2 | 18 | Kamphaeng Phet, Nakhon Sawan, Phichit, Uthai Thani |
| | Upper Central Region Area 1 | 2 | Chai Nat |
| 4 | Upper Central Region Area 1 | 1 | Nonthaburi, Pathum Thani, Phra Nakhon Si Ayutthaya, Saraburi |
| | Upper Central Region Area 2 | 2 | Lopburi, Sing Buri, Ang Thong |
| | Middle Central Region Area | 3 | Nakhon Nayok |
| 5 | Lower Central Region Area 1 | 4 | Kanchanaburi, Nakhon Pathom, Ratchaburi, Suphan Buri |
| | Lower Central Region Area 2 | 5 | Prachuap Khiri Khan, Phetchaburi, Samut Songkhram, Samut Sakhon |
| 6 | Middle Central Region Area | 3 | Prachinburi, Sa Kaeo |
| | Eastern Region | 9 | Chanthaburi, Trat, Rayong, Chonburi, Samut Prakan, Chachoengsao |
| 7 | Middle Northeastern Region Area | 12 | Kalasin, Khon Kaen, Maha Sarakham, Roi Et |
| 8 | Upper Northeastern Region Area 1 | 10 | Loei, Nong Khai, Nong Bua Lamphu, Udon Thani, Bueng Kan |
| | Upper Northeastern Region Area 2 | 11 | Nakhon Phanom, Mukdahan, Sakon Nakhon |
| 9 | Lower Northeastern Region Area 1 | 13 | Chaiyaphum, Nakhon Ratchasima, Buriram, Surin |
| 10 | Lower Northeastern Region Area 2 | 14 | Yasothon, Sisaket, Amnat Charoen, Ubon Ratchathani |
| 11 | Southern Region Eastern Seaboard | 6 | Chumphon, Surat Thani, Nakhon Si Thammarat |
| | Southern Region Western Seaboard | 7 | Ranong, Phang Nga, Phuket, Krabi |
| 12 | Southern Region Eastern Seaboard | 6 | Phatthalung |
| | Southern Region Western Seaboard | 7 | Trang |
| | Southern Region Border Provinces | 8 | Songkhla, Satun, Pattani, Yala, Narathiwat |

Table S2 List of parameter values applied in the study.

| Parameter | Value (min, max) | Reference |
|---|---|---|
| **Total population** | Bangkok: 8300000<br>Health District 1: 5900000<br>Health District 2: 3500000<br>Health District 3: 3000000<br>Health District 4: 5300000<br>Health District 5: 4200000<br>Health District 6: 5900000<br>Health District 7: 5000000<br>Health District 8: 5500000<br>Health District 9: 6700000<br>Health District 10: 4500000<br>Health District 11: 4500000<br>Health District 12: 4900000 | (8,14) |
| **Initial infectious population** | 10 | (20) |
| **Infection rate (Ki)**<br>**Contact rate * transmission probability** | Fit to each health district's data | |
| **Start day of simulation** | 01.02.2020 | |
| **Time to infectious (days)** | (3.4, 4.5) | (11) |
| **Time to symptomatic (days)** | (2.4, 3.5) | (20,21) |
| **Time to hospitalization (days)** | (3, 6) | (20) |
| **Time to critical (days)** | (4, 6) | (20) |
| **Time to death (days)** | (4, 6) | (20) |
| **Time to detection** | (2, 2) | (20,22) |
| **Time to detection As** | (1, 6) | |
| **Time to detection Sym** | (7, 7) | |
| **Time to detection Sys** | (2, 2) | |
| **Recovery time to asymp** | (7, 10) | |
| **Recovery time asymptomatic (days)** | (7, 10) | (20,23) |
| **Recovery time mild symptomatic (days)** | (9, 9) | Assumed to be same as recovery rate in asymptomatics |
| **Recovery time hospitalized (days)** | (4, 6) | (20) |
| **Recovery time critical (days)** | (8, 10) | (20) |
| **Fraction symptomatic** | (0.5, 0.7) | (24) |
| **Fraction severe symptomatic** | (0.2,0.31) | (25,26) |
| **Fraction critical** | (0.2, 0.35) | (27) |
| **Case fatality rate** | (0.02,0.1) | (28,29) |
| **Reduced infectiousness of detected cases** | (0, 0.3) | (20) |
| **Social multiplier 1-11** | Fitted to Thailand's data | |

We applied transmission reductions derived from containment measures in separate events based on the timeline of events was based on Thailand's government containment measures (Table S3) and mobility data (Figure S1). The specified dates are in the middle of the week to reduce possible fluctuation during weekends.

Table S3 Thai government's containment measures timeline.

| **Containment measures** | **Affected area** | **Start date of implementation** |
|---|---|---|
| The Notification of the Civil Aviation Authority of Thailand on Practical Guideline for Air Operators Performing Flights into the Kingdom of Thailand. Passengers traveling to the Kingdom of Thailand are required to perform the screening of passengers at the time of check-in. | Air travels | 22.03.2020 |
| Declared a state of emergency under the Emergency Decree on Public Administration in 2548. | Nationwide | 26.03.2020 |
| 7 Decree requirements. Emergency. No gathering in public spaces and closing borders | Nationwide | 26.03.2020 |
| Curfew (22:00-04:00) declared a state of emergency under the Emergency Decree 2548 No. 2. | Nationwide | 03.04.2020 |
| 5 easing phases | Nationwide | Phase I: 03.05.2020<br>Phase II: 17.05.2020<br>Phase III: 01.06.2020<br>Phase IV: 15.06.2020<br>Phase V: 29.06.2020 |

Figure S1 Thailand's mobility data and timeline of transmission reduction events applied in the model.

Figure S1 shows changes in mobility of Thai population by category of locations compared to baseline. The baseline day represents a normal value for that day of the week taken from the median value from the 5-week period of Jan 3 to Feb 6, 2020. The figure was produced from Thailand data of Google's COVID-19 community mobility report which came from aggregated, anonymized insights the company uses in products such as Google Maps. The report provides movement trends over time by geography, across different categories of places.

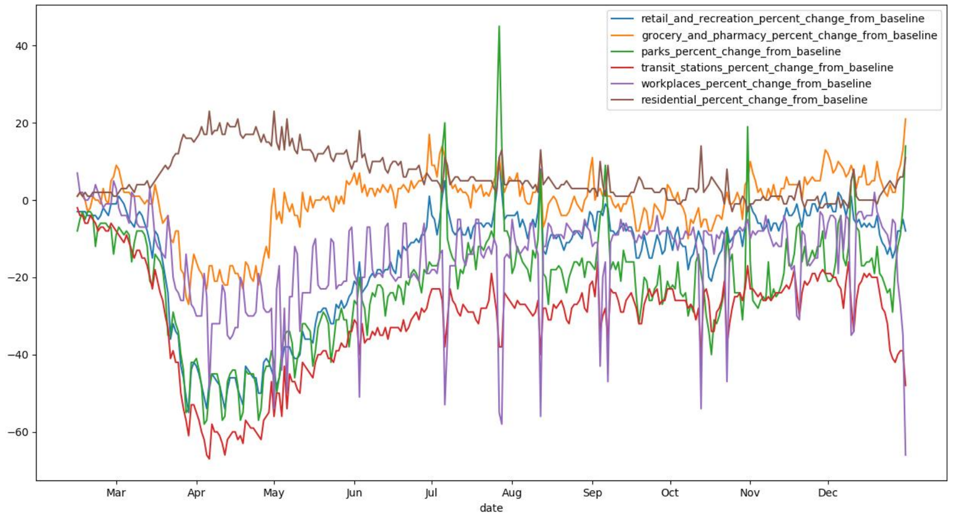

Table S4 Dates of changes in transmission (grey areas mean not applicable).

| Health District Number | Date | | | | | | | | | | |
|---|---|---|---|---|---|---|---|---|---|---|---|
| | 1st | 2nd | 3rd | 4th | 5th | 6th | 7th | 8th | 9th | 10th | 11th |
| 1 | Mar 12 | Mar 17 | Mar 21 | Apr 21 | Jul 15 | Oct 15 | Nov 25 | | | | |
| 2 | Mar 15 | Mar 17 | Mar 21 | Apr 21 | Jul 15 | Aug 25 | Oct 15 | Dec 20 | Dec 21 | Dec 25 | Dec 28 |
| 3 | Mar 10 | Mar 17 | Mar 21 | Apr 21 | Jul 15 | Oct 15 | Nov 20 | | | | |
| 4 | Mar 12 | Mar 17 | Mar 21 | Apr 21 | Jul 15 | Oct 1 | Dec 20 | | | | |
| 5 | Mar 12 | Mar 17 | Mar 21 | Apr 21 | Jul 15 | Oct 30 | Dec 1 | | | | |
| 6 | Mar 12 | Mar 17 | Mar 21 | Apr 21 | Jul 15 | Oct 17 | Dec 15 | | | | |
| 7 | Mar 12 | Mar 17 | Mar 21 | Apr 21 | Jul 15 | Oct 1 | Nov 1 | | | | |
| 8 | Mar 12 | Mar 17 | Mar 21 | Apr 21 | Jul 15 | Nov 25 | Dec 5 | | | | |
| 9 | Mar 12 | Mar 17 | Mar 21 | Apr 21 | Jul 15 | Oct 1 | Dec 20 | | | | |
| 10 | Mar 12 | Mar 17 | Mar 21 | Apr 21 | Jul 15 | Nov 25 | Dec 5 | | | | |
| 11 | Mar 12 | Mar 17 | Mar 21 | Apr 21 | Jul 15 | Oct 5 | Nov 5 | | | | |
| 12 | Mar 12 | Mar 17 | Mar 21 | Apr 21 | Jul 15 | Nov 15 | Dec 15 | | | | |
| 13 | Mar 23 | Mar 24 | Mar 25 | Apr 21 | Jul 15 | Oct 15 | Dec 30 | | | | |

Table S5 The range of changes in transmission applied in the study.

| Health District | Ki | Multiplier 1 | Multiplier 2 | Multiplier 3 | Multiplier 4 | Multiplier 5 | Multiplier 6 | Multiplier 7 | Multiplier 8 | Multiplier 9 | Multiplier 10 | Multiplier 11 |
|---|---|---|---|---|---|---|---|---|---|---|---|---|
| 1 | (0.235,0.25) | (2.5, 2.7) | (1.5,1.7) | (0.245, 0.251) | (0.13,0.23) | (1.359, 1.364) | (1.55,1.65) | (0.2,0.3) | | | | |
| 2 | (0.325,0.375) | (3.493,3.514) | (2.253, 2.288) | (0.25,0.03) | (0.13,0.23) | (1.35,1.364) | (0.6,0.7) | (0.13,0.23) | (2,2.5) | (0.013, 0.023) | (6,6.5) | (0.3,0.5) |
| 3 | (0.291,0.294) | (3.132,3.211) | (2.983, 3.056) | (0.245, 0.255) | (0.13,0.23) | (1.328, 1.364) | (2.038, 3) | (0.13,0.23) | | | | |
| 4 | (0.233,0.239) | (2,2.3) | (2,2) | (0.245, 0.263) | (0.13,0.23) | (1.324, 1.357) | (0.95,1.05) | (0.13,0.23) | | | | |
| 5 | (0.265,0.273) | (1.279,1.404) | (0.9,1.1) | (0.239, 0.258) | (0.13,0.23) | (1.347, 1.385) | (3.75,4) | (0.13,0.23) | | | | |
| 6 | (0.265,0.273) | (2.3, 2.5) | (0.239, 0.258) | (0.239, 0.258) | (0.13,0.23) | (1.347, 1.385) | (1.28,1.32) | (0.13,0.23) | | | | |
| 7 | (0.265,0.271) | (3.5, 3.7) | (2.294, 2.356) | (0.026, 0.035) | (0.13,0.23) | (1.333, 1.373) | (1.314, 1.368) | (0.373, 0.409) | | | | |
| 8 | (0.28,0.3) | (3,3.2) | (2.876, 2.928) | (0.245, 0.263) | (0.13,0.23) | (1.324, 1.357) | (9.5,10.5) | (0.13,0.23) | | | | |
| 9 | (0.27,0.3) | (4.3, 4.8) | (1.5,2) | (0.025, 0.03) | (0.13,0.23) | (1.324, 1.357) | (0.8,0.9) | (0.13,0.23) | | | | |
| 10 | (0.297,0.304) | (3,3.1) | (2.876, 2.928) | (0.097, 0.102) | (0.13,0.23) | (1.324, 1.357) | (8.3,9.3) | (0.13,0.23) | | | | |
| 11 | (0.328,0.34) | (0.4, 0.6) | (0.2,0.3) | (0.242, 0.258) | (0.13,0.23) | (1.309, 1.359) | (1.2,1.3) | (0.13,0.23) | | | | |
| 12 | (0.34,0.345) | (0.7, 0.9) | (0.25,0.35) | (0.245, 0.259) | (0.13,0.23) | (1.339, 1.366) | (1.01,1.15) | (0.13,0.23) | | | | |
| 13 | (0.24,0.26) | (0.0001,0.0003) | (0.0001,0.0005) | (0.0001,0.0003) | (0.13,0.23) | (1.34,1.36) | (1.15,1.25) | (0.23,0.33) | | | | |
